# S-ketamine in patient-controlled analgesia reduces opioid consumption in a dose-dependent manner after major lumbar fusion surgery: a randomized, double-blind, placebo-controlled clinical trial

**DOI:** 10.1101/2021.01.22.21250352

**Authors:** Elina C.V. Brinck, Taru Virtanen, Sanna Mäkelä, Venla Soini, Ville-Veikko Hynninen, Jukka Mulo, Urmas Savolainen, Juho Rantakokko, Kreu Maisniemi, Antti Liukas, Klaus T. Olkkola, Vesa Kontinen, Pekka Tarkkila, Marko Peltoniemi, Teijo I. Saari

## Abstract

**BACKGROUND:** Spinal fusion surgery causes severe pain. Strong opioids, commonly used as postoperative analgesics, may have unwanted side effects. S-ketamine may be an effective analgesic adjuvant in opioid patient-controlled analgesia (PCA). However, the optimal adjunct S-ketamine dose to reduce postoperative opioid consumption is still unknown.

**METHODS:** We randomized 107 patients at two tertiary hospitals in a double-blinded, placebo-controlled clinical trial of adults undergoing major lumbar spinal fusion surgery. Patients were randomly allocated to four groups in order to compare the effects of three different doses of adjunct S-ketamine (0.25, 0.5, and 0.75 mg ml^−1^) or placebo on postoperative analgesia in oxycodone PCA. Study drugs were administered for 24 hours postoperative after which oxycodone-PCA was continued for further 48 hours. Our primary outcome was cumulative oxycodone consumption at 24 hours after surgery.

**RESULTS:** Of the 100 patients analyzed, patients receiving 0.75 mg ml^−1^ S-ketamine in oxycodone PCA needed 25% less oxycodone at 24 h postoperatively (61.2 mg) compared with patients receiving 0.5 mg ml^−1^ (74.7 mg) or 0.25 mg ml^−1^ (74.1 mg) S-ketamine in oxycodone or oxycodone alone (81.9 mg) (mean difference: −20.6 mg; 95% confidence interval [CI]: −41 to −0.20; *P* = 0.048). A beneficial effect in mean change of pain intensity at rest was seen in the group receiving 0.75 mg ml^−1^ S-ketamine in oxycodone PCA compared with patients receiving lower ketamine doses or oxycodone alone (standardized effect size: 0.17, 95% CI: 0.013–0.32, *P =* 0.033). The occurrence of adverse events was similar among the groups.

**CONCLUSIONS:** Oxycodone PCA containing S-ketamine as an adjunct at a ratio of 1: 0.75 decreased cumulative oxycodone consumption at 24 h after major lumbar spinal fusion surgery without additional adverse effects.

## Introduction

Severe postoperative pain is common after major spinal fusion surgery [1]. Inadequate pain management is associated with increased postoperative complications, delayed ambulation, and chronic postoperative pain [2]. This may lead to unplanned readmission after surgery, decreased patient satisfaction, and increased health-care costs. Opioids have been the cornerstone of postoperative analgesia after major surgery, but the increasing awareness of the problems associated with their use, such as opioid-associated adverse effects, opioid-induced hyperalgesia, and the risk of addiction, combined with the current opioid crisis has encouraged the search for alternative analgesic strategies.

Multimodal analgesia targets different pain signaling pathways by combining two or more analgesic modalities, aiming at additive or even synergistic analgesic effect [3]. Multimodal analgesia has proven feasible after major spinal fusion surgery in an effort to optimize pain relief while minimizing opioid-related adverse effects [4,5].

Patient-controlled analgesia (PCA) use is associated with lower pain intensity and greater patient satisfaction compared with conventional (oral, subcutaneous, or intramuscular) administration routes [6]. PCA may enhance patient autonomy because the analgesic drug is readily available. Morphine is the most frequently used opioid in PCA, while oxycodone use is associated with higher patient satisfaction scores [6,7].

Low-dose (<1 mg kg^−1^) ketamine inhibits N-methyl-D-aspartate receptors in nociceptive neurons and activates descending inhibitory pain pathways, resulting in attenuated wind up and central sensitization [8–11]. The effect of perioperative intravenous ketamine as an adjunct analgesic has been documented in several clinical trials and meta-analyses [12]. A recent review article concluded that combining ketamine with an opioid in IV-PCA during the postoperative period has a beneficial effect on analgesia and opioid consumption [13]. In orthopedic surgery, earlier trials found negative or unclear results with opioid-ketamine PCA [14–16]. However, studies’ vast heterogeneity and small sample sizes have failed to establish a possible a possible dose-responsiveness. Thus, the optimal ketamine to opioid ratio in intravenous PCA is yet to be elucidated.

We hypothesized that adjunct S-ketamine with oxycodone in an intravenous PCA is superior to oxycodone, with reduced opioid-associated adverse events in the immediate postoperative period after major spinal surgery. This randomized, double-blind, placebo-controlled study compared three different doses of S-ketamine with placebo added as an adjuvant to oxycodone IV-PCA administered for 24 hours after lumbar spinal fusion in adult patients. After the study phase, IV oxycodone-PCA without ketamine was continued for further 48 hours. The primary outcome was cumulative oxycodone consumption at 24 h after surgery. The secondary endpoints included postoperative pain intensity, oxycodone consumption, and occurrence of adverse events up to 72 h after surgery.

## Methods

### Ethics and registration

This study (DoseRespKeta) was approved by the institutional review board of the Hospital District of Southwest Finland (number: *103/1800/2016*) and the Finnish Medicines Agency (FIMEA, KL 135/2016). The trial was registered before patient enrollment at clinicaltrials.gov (NCT02994173; Principal investigator: TIS; Date of registration: September 11, 2017) and in the EudraCT database (*2016-002887-14*; Principal investigator: TIS; Date of registration: October 12, 2016). No changes to methods were made after trial commencement. The detailed study protocol is available upon request from the authors. Written informed consent was obtained from the participants before inclusion to the study. This manuscript complies with the Consolidated Standards of Reporting Trials guidelines.

### Patient population

This two-center study was carried out at the T-hospital and TYKS ORTO units at Turku University Hospital in Turku, Southwest Finland and the HUS Helsinki University Hospital’s Töölö Unit in Helsinki, Finland. One hundred and seven adult patients scheduled for elective posterolateral lumbar spine fusion with bilateral transpedicular screw instrumentation under general anesthesia were recruited between 6 February 2017 and 31 October 2019 (Fig 1).

**Figure 1.**
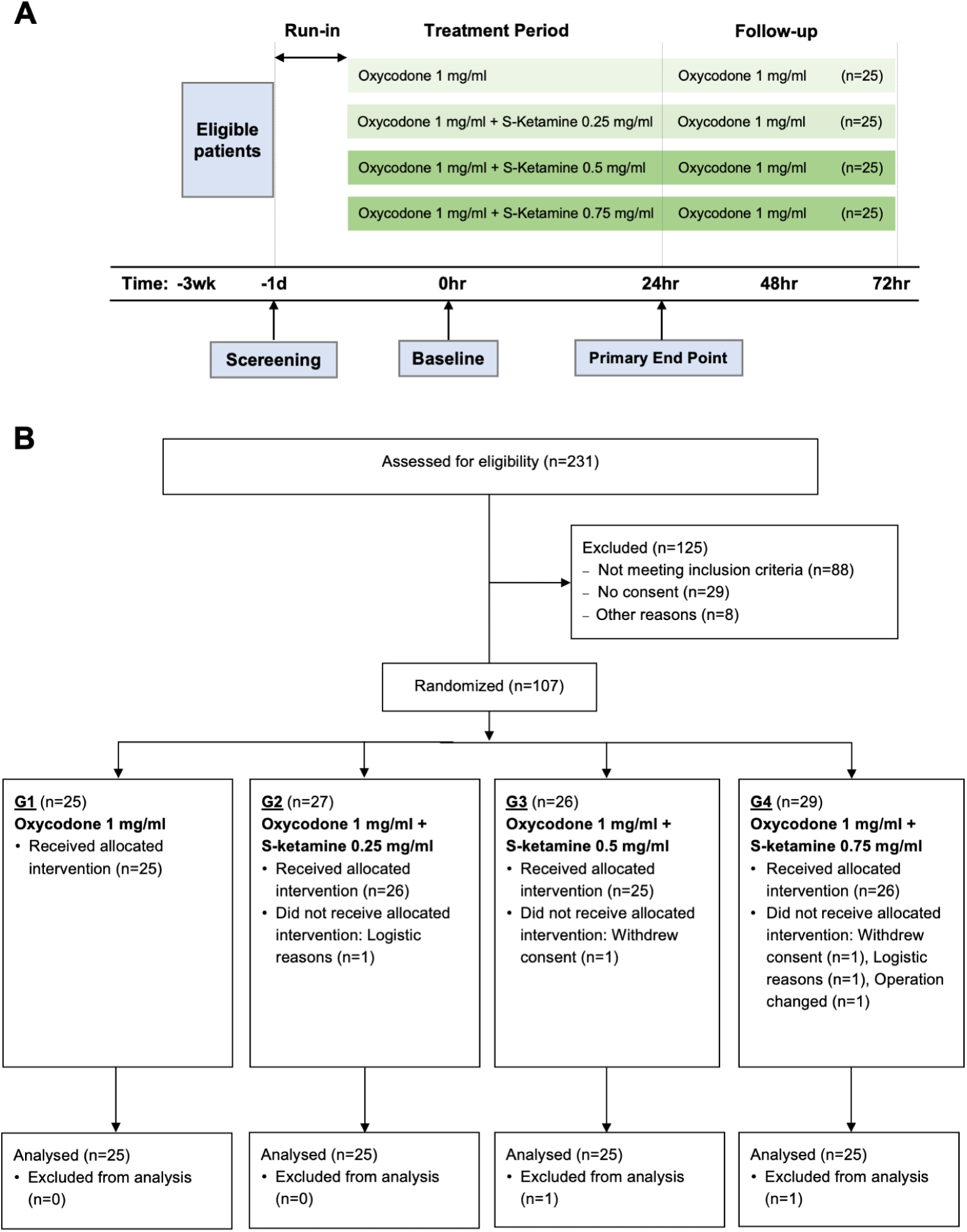
DoseRespKeta trial structure and CONSORT 2009 flow diagram. CONSORT indicates Consolidated Standards of Reporting Trials.

Patients with previous history of intolerance to a study drug, concomitant drug therapy with strong opioids or strong cytochrome P450 3A4 or 2B6 inductor or inhibitors 2 weeks prior to study, younger than 20 years and older than 80 years, BMI >35, sleep apnea or any other sleep disorder, existing significant liver or kidney disease, history of alcoholism, drug abuse, and psychological or other emotional problems likely to invalidate informed consent were excluded. Patients were pre-screened by a preoperative care nurse. Potentially eligible subjects were directed to investigators for further screening and information. ECVB and MP enrolled all participants.

### Study design, randomization, and blinding

A randomized, double-blind, controlled dose-response study design was used (Fig 1). The allocation ratio was 1:1:1:1. Subjects were randomly assigned to one of four dosing groups (G) for intravenous PCA:

- G1, oxycodone 1 mg ml^−1^ alone
- G2, oxycodone 1 mg ml^−1^ + S-ketamine 0.25 mg ml^−1^ (ratio 1:0.25)
- G3, oxycodone 1 mg ml^−1^ + S-ketamine 0.5 mg ml^−1^ (ratio 1:0.5),
- G4, oxycodone 1 mg ml^−1^ + S-ketamine 0.75 mg ml^−1^ (ratio 1:0.75).

An independent statistician created a computer-generated randomization list. The list was sent to the local hospital pharmacy, which took care of assignment. Coded PCA reservoirs with no other markings were delivered to the operation room by the pharmacy on the day of surgery to ensure double blinding. Patients, researchers, and clinical staff were blinded to group allocation.

### Conduct of the study and anesthesia

After providing written informed consent, patients learned to use the PCA system (CADD®-Solis VIP and CADD-Legacy® PCA Pump Model 6300, Smiths Medical, Minneapolis, MN, USA) and numerical rating scale (NRS; 0 denotes no pain, and 10 indicates the worst imaginable pain). Preoperatively, all patients received 1000 mg of paracetamol orally. Routine anesthetic monitoring included pulse oximetry, electrocardiography, invasive blood pressure via radial artery cannulation, and body temperature measurement. General anesthesia was induced and maintained with propofol (Propofol-Lipuro 20 mg ml^−1^; B. Braun Melsungen AG, Melsungen, Germany) and remifentanil (Remifentanil B. Braun 1 mg ml^−1^; B. Braun Melsungen AG, Melsungen, Germany) target-controlled infusions. Schnider- and Minto-effect-site models were used for propofol and remifentanil, respectively [17,18]. No opioids other than remifentanil were used before or during anesthesia. Rocuronium bromide (Esmeron 10 mg ml^.-1^; Merck Sharp & Dohme B.V., BN Haarlem, The Netherlands) 0.4–0.6 mg kg^−1^ facilitated endotracheal intubation. The depth of anesthesia (bispectral index BIS or entropy index) was targeted from 45 to 55 with entropy (GE B850 Monitor Entropy Module, GE, Helsinki, Finland) or BIS monitor (The Philips BIS module, Philips Medical Systems, Eindhoven, The Netherlands). Mean arterial pressure (MAP) was maintained at 65–75 mmHg. Intravenous bolus doses of ephedrine and/or noradrenaline infusion were administered if necessary. Local anesthetic was injected to the skin incision area before incision (lidocaine 5 mg kg^−1^ c. adrenaline 5 µg kg^−1^; Orion Pharma, Espoo, Finland) and after wound closure (levobupivacaine 2.5 mg kg^−1^; Chirocaine 2.5 mg ml^.1^, AbbVie S.r.l., Campoverde di Aprilia, Italy) as per hospital routines.

At the end of surgery, the PCA pump was attached to the intravenous line and activated. The first dose of PCA solution was given just after the cessation of propofol and remifentanil infusions, before patients woke up from anesthesia. As soon as the patient awoke, they were encouraged to use PCA to treat postoperative pain if necessary. The starting dose of oxycodone (Oxycodone Orion 10 mg ml^−1^; Orion Pharma, Espoo, Finland) in the PCA solution was 2 mg with a lockout interval of 5 min. When NRS was 4 or lower, the PCA oxycodone dose was decreased to 1 mg (G1–G4). The study PCA dosing continued for 24 h from the end of surgery, after which the PCA cassettes were changed in all study groups and contained only 1 mg/ml oxycodone thereafter in all study groups.

The total duration of PCA treatment was three days. Postoperative nausea and vomiting (PONV) were treated with intravenous ondansetron and dehydrobenzperidol if necessary. Intravenous bolus of lorazepam (1mg) was administered if a patient had unpleasant psychomimetic symptoms.

### Measurements and data handling

Heart rate, blood pressure, respiratory rate, peripheral oxygen saturation, NRS (0–10) for pain intensity at rest and upon movement, level of sedation with the Richmond Agitation-Sedation scale (RASS), nausea, vomiting, pruritus, unpleasant dreams or any other adverse effect or sensation thought to be caused by PCA, cumulative PCA pressings, or the doses given were registered immediately after arrival in the recovery room and at the following time points: five, 30, 60, 120, and 240 minutes and eight, 24, 48, and 72 hours later. Patients evaluated their pain relief satisfaction (yes/no) at the end of postoperative days 1–3.

All clinical patient data were collected on individual case report forms. All data were subsequently transferred to electronic format for exploratory data analysis.

### Primary and secondary outcomes

The primary endpoint was cumulative oxycodone consumption at 24 h after surgery, the end of period when three different ratios of S-ketamine were added to oxycodone PCA solution.

Secondary outcomes were NRS and RASS ratings, oxycodone consumption, PONV, pruritus, and unpleasant dreams or other adverse effects.

### Sample size calculation

Based on a previous study [19], we calculated that 25 patients per group were necessary to demonstrate a clinically significant 25% reduction in oxycodone consumption with a level of significance of *P =* 0.05 and power of 80%.

### Statistical analyses

The authors approved the statistical analysis plan before the analyses began. Explorative data analysis was conducted before statistical inference by plotting and tabulating the data. Normality assumptions were tested before analysis, using probit plots and the Shapiro–Wilk W-test. Levene’s test was used to evaluate homogeneity of variances.

The primary outcome measure, cumulative opioid consumption during the first 24 h is presented as both median and interquartile range (IQR; Q1–Q3) and mean (SD). Cumulative opioid consumption at 24, 48, and 72 h was analyzed by using a linear mixed model for repeated measurements. The model included the following factors: PCA dosing groups G1–G4, age (as a continuous covariate), age, gender, prior use of opioids (yes/no), prior use of gabapentinoids (yes/no), cumulative amount of dose requests during PCA, and postoperative time (24, 48, and 72 h). In addition, the interaction with time and opioid consumption and prior use of weak opioids was examined. Non-significant terms were excluded from the final model. An unstructured covariance structure was used. Square-root transformation was used to fulfill assumptions of the model. Studentized residuals were used to check assumptions.

To study pain burden, we analyzed if the mean change in NRS over time differed between the groups during the first 24 hours postoperatively. A hierarchical linear mixed model was used including time as a within-factor (with five time points), PCA dosing group as a between-factor, and their interaction. The time factor was handled as categorical to estimate all possible shapes of mean changes over time. NRS values were standardized before analysis by centering and scaling to have a mean of zero and a SD of one. Effect sizes were computed as the eta squared based on the H-statistic [20]. We also studied in a separate model, whether age, sex, weight, chronic pain, and prior use of weak opioids or gabapentinoids affected the results. Differences in NRS and sedation (measured with RASS) between the groups were evaluated at the end of post-anesthesia care unit (PACU) treatment (t = 480 min) and at 24 h with a Kruskal– Wallis test.

Differences in postoperative nausea and vomiting, pruritus, and unpleasant dreams (yes/no) among groups at 24, 48, and 72 h postoperatively were evaluated with ordinal logistic regression. The effect of age, sex, weight, chronic pain, and prior use of weak opioids or gabapentinoids on the results were tested as covariate effects. To analyze the effect of adjuvant ketamine treatment on these parameters, we included changes in parameters to the model during the previous 24 h.

Descriptive statistics are shown as means and SD when variables are normally distributed and otherwise as medians and IRQs. Categorical variables are summarized as counts and percentages. The statistical significance level was set to *P* <0.05. RStudio (version 1.0.153) [21] with R (version 3.6.0) [22] and ggplot2 (version 2.2.1) were used for statistical analyses and graphical presentations.

## Results

A total of 231 patients were assessed for eligibility. 107 patients were recruited in the study (Fig 1B) between February 2017 and October 2019. Two patients withdrew their consent before randomization and were excluded. Two patients were further excluded due to logistical reasons before surgery. In one case, the operation plan was changed after randomization and this patient was also excluded from the study. Two more patients withdrew consent during the first 24 h after the start of intervention and were excluded from the final analysis. Thus, 25 patients in each of the four groups were included in the analysis. Patient median age was 60 (28–78) years and 77% were women. No significant differences in baselines characteristics were present among the groups (Table 1).

**Table 1.** Perioperative Data and Characteristics of Patients Who Were Included in the Analysis.

|  | All | G1 | G2 | SMD<br>Between<br>G1 and G2 | G3 | SMD<br>Between<br>G1 and G3 | G4 | SMD<br>Between<br>G1 and G4 |
| --- | --- | --- | --- | --- | --- | --- | --- | --- |
| n | 100 | 25 | 25 |  | 25 |  | 25 |  |
| Age, yr (median, range) | 60 (28-78)<sup>2</sup> | 56 (34-78) | 62 (28-76) | 0.214 | 60 (40-75) | 0.433 | 62 (30-76) | 0.329 |
| Female/male, n (%) | 45/55 | 11/14 (44%)<sup>1</sup> | 11/14 (44%) | <0.001 | 12/13 (48%) | <0.001 | 11/14 (44%) | 0.08 |
| Body weight (kg) | 77.7 (15.6) | 77.7 (15.6) | 74.9 (15.9) | 0.215 | 79.0 (14.1) | 0.278 | 77.0 (11.2) | 0.156 |
| BMI (kg/m<sup>2</sup>) | 26.9 (4.3) | 26.6 (4.6) | 27.3 (4.3) | 0.003 | 26.9 (3.5) | 0.152 | 26.8 (4.9) | 0.063 |
| Smoking (yes/no), n (%) | 9/91 | 1/24 (4%)<sup>1</sup> | 3/22 (12%) | 0.169 | 3/22 (12%) | 0.298 | 2/23 (7.7%) | 0.298 |
| Previous chronic pain (yes/no), n (%) | 15/85 | 22/3 (88%)<sup>1</sup> | 19/6 (76%) | 0.115 | 22/3 (88%) | 0.316 | 22/3 (88%) | <0.001 |
| Median [IRQ] preoperative pain intensity (NRS) | 6 [4, 8] | 6 [5, 7] | 5 [5, 7] | 0.1 | 7 [5, 8] | 0.42 | 7 [5, 8] | 0.023 |
| Preoperative opioid (yes/no), n (%) | 5/94 | 2/23 (8.0%)<sup>1</sup> | 2/23 (8.0%) | 0.169 | 0/25 (0%) | <0.001 | 1/24 (3.8%) | 0.417 |
| Preoperative gabapentinod (yes/no), n (%) | 18/82 | 3/22 (12%)<sup>1</sup> | 3/22 (12%) | 0.22 | 7/18 (28%) | <0.001 | 5/20 (19%) | 0.408 |
| Diabetes (yes/no), n (%) | 8/92 | 1/24 (4.0%)<sup>1</sup> | 3/22 (12%) | 0.169 | 1/25 (4.0%) | 0.298 | 3/22 (11%) | <0.001 |
| ASA physical status, n (%) |  |  |  | 0.408 |  | 0.182 |  | 0.331 |
| 1 | 24 | 7 (28.0) | 6 (24.0) |  | 6 (24.0) |  | 5 (20.0) |  |
| 2 | 66 | 17 (68.0) | 15 (60.0) |  | 17 (68.0) |  | 17 (68.0) |  |
| 3 | 10 | 1 (4.0) | 4 (16.0) |  | 2 (8.0) |  | 3 (12.0) |  |
| Duration of surgery (min) | 306 (94) | 325 (89) | 292 (64) | 0.424 | 303 (125) | 0.202 | 305 (89) | 0.226 |
| Propofol during surgery (mg·kg<sup>-1</sup>) | 37 (12) | 39 (11) | 36 (8) | 0.259 | 35 (17) | 0.26 | 36 (10) | 0.248 |
| Remifentanyl during surgery (µg·kg<sup>-1</sup>·min<sup>-1</sup>) | 0.12 (0.05) | 0.13 (0.06) | 0.12 (0.04) | 0.27 | 0.11 (0.04) | 0.429 | 0.11 (0.04) | 0.383 |

Data are mean (standard deviation) or [range) unless otherwise stated. Standardized difference is the difference in the mean of a variable between 2 groups divided by an estimate of the standard deviation of that variable [22].
G1, Patient controlled intravenous oxycodone 1 mg/ml (n=25); G2, Patient controlled intravenous oxycodone 1 mg/ml+ ketamine 0.25 mg/ml (n=25); G3, Patient controlled intravenous oxycodone 1 mg/ml+ ketamine 0.5 mg/ml (n=25); G4, Patient controlled intravenous oxycodone 1 mg/ml+ ketamine 0.75 mg/ml (n=25); SMD, standardized mean deviation; n, number of non-missing value; BMI, body-mass index; IQR, interquartile range; NRS, numerical rating scale; ASA, American Society of Anesthesiologists.
<sup>1</sup>Percentage of females or yes-answers

### Opioid consumption

Cumulative oxycodone consumption was highest in group G1 and lowest in G4 (Fig 2, Table 2). The median total oxycodone consumption during the first 24 h after surgery was 81.9 mg (IQR: 63.2–101), 74.1 mg (IQR: 62.1–86.1), 74.7 mg (IQR: 62.2–87.1), and 61.3 mg (IQR: 48.7–73.8) in groups G1–G4, respectively. Postoperative cumulative oxycodone consumption was significantly reduced in the group with the highest ketamine concentration (G4) compared with the control group (G1): the mean difference was −21 mg (95% CI: −41 to −0.2, *P =* 0.048), −26 mg (95% CI: −55 to −6.2, *P =* 0.044) and −41 mg (95% CI: −68 to −14, *P =* 0.003) at 24, 48, and 72 h after surgery, respectively (Fig 2). There was no significant difference in time to dose reduction from 2 mg to 1 mg PCA-bolus among groups (median [IQR]: 125 min [78–203], 127 min [82– 174], 112 min [69–185] and 145 min [79–211] in groups G1–G4, respectively, *P =* 0.838).

**Table 2.**
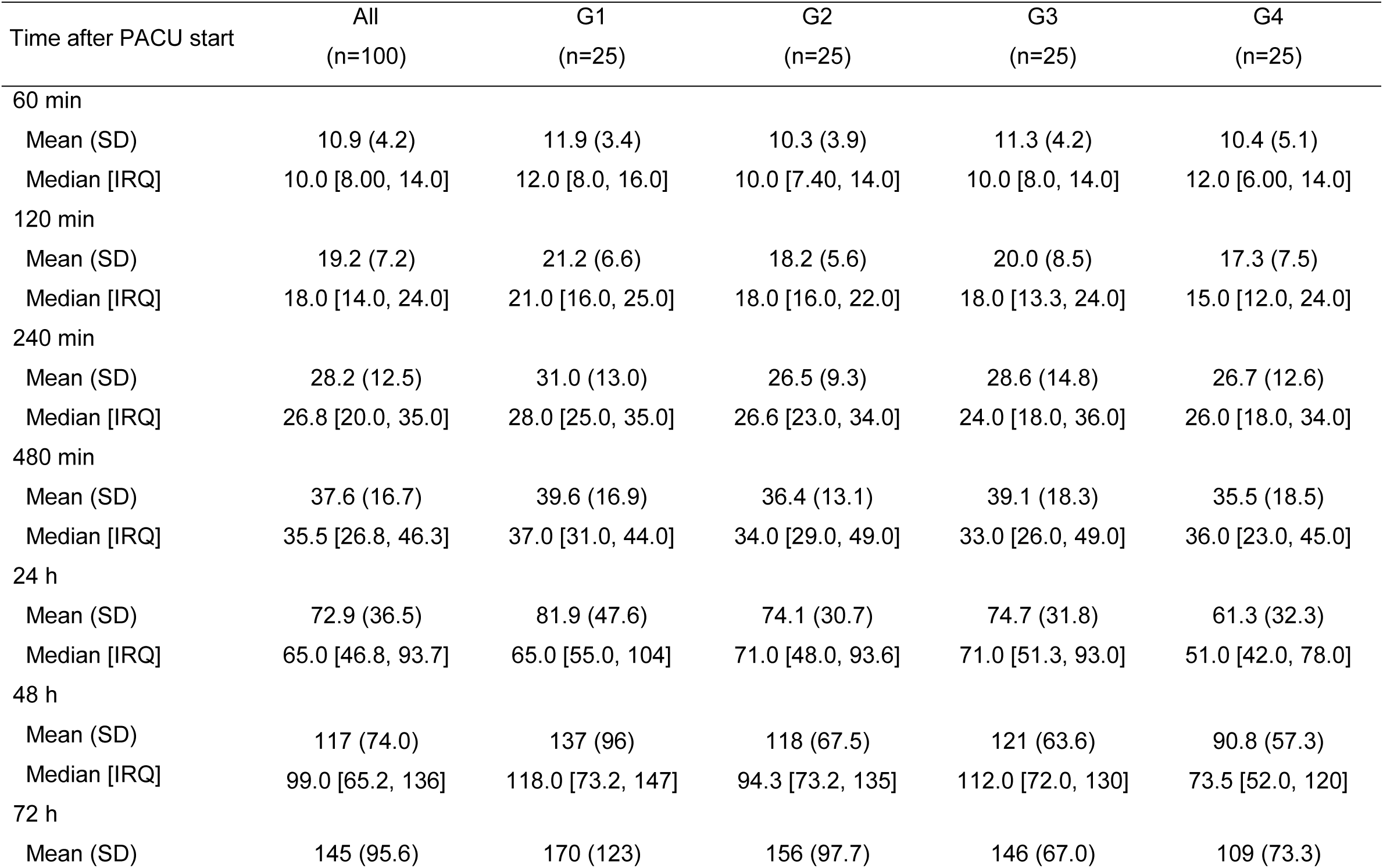
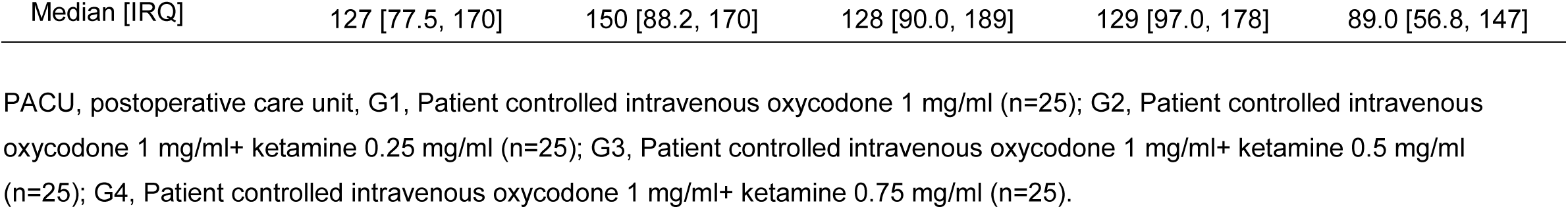
Oxycodone consumption during the study after the beginning of PACU care

**Figure 2.**
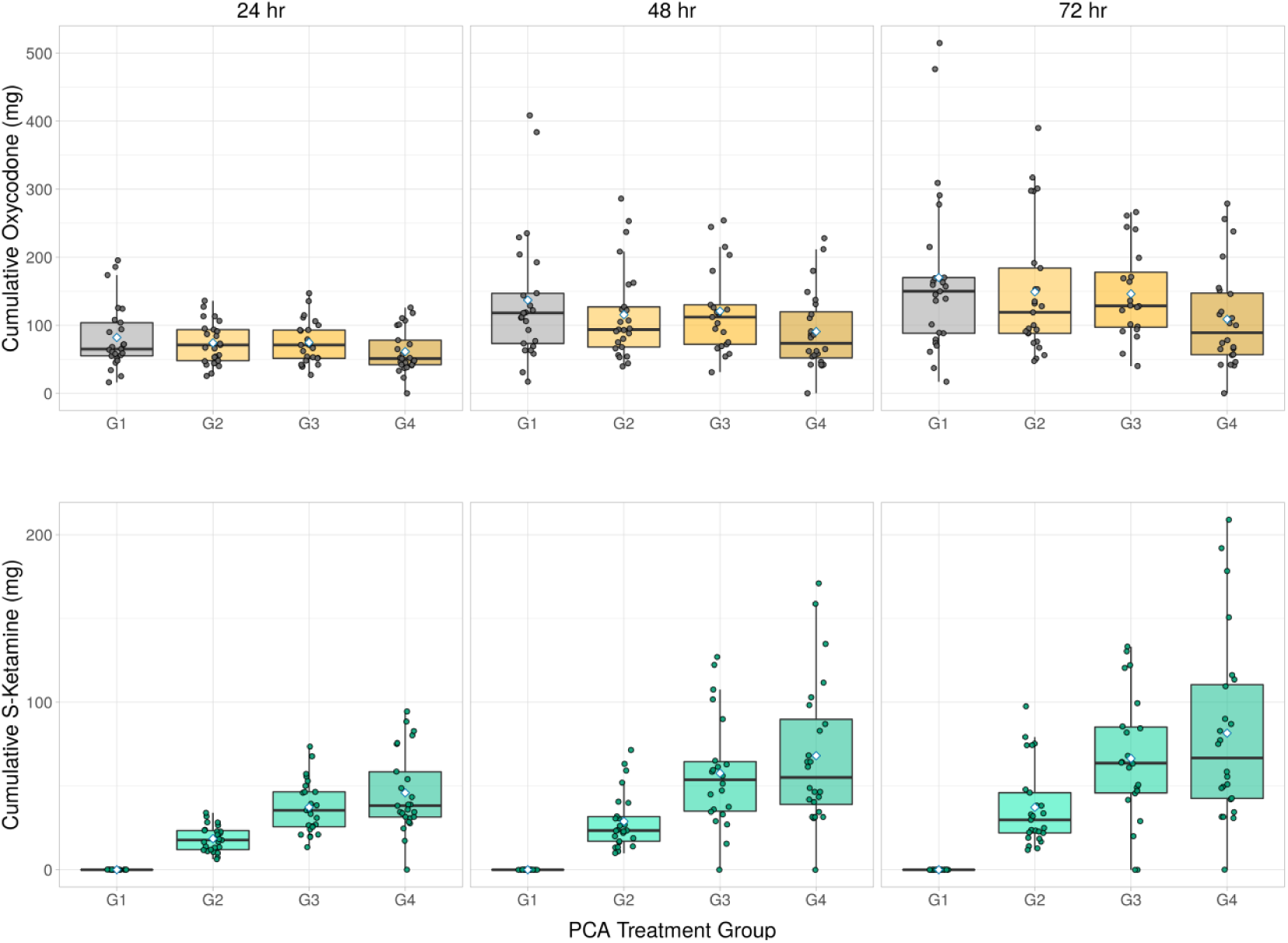
Postoperative cumulative oxycodone (*top row*) and S-ketamine (*bottom row*) consumption during the first 24 h in four patient-controlled analgesia (PCA) treatment groups (G1¬ –G4). PCA, patient-controlled analgesia. Box plots show the median and 25–75th percentiles, and the whiskers indicate the minimum and maximum. Blue diamonds show the mean oxycodone consumption in each plot.

### Cumulative S-ketamine consumption at 24, 48 and 72 h

Cumulative S-ketamine consumption was highest in group G1 and lowest in G4 (Fig 2b). The median total S-ketamine consumption during the first 24 h after surgery was 17.8 mg (IQR: 12.0–23.4), 35.5 mg (IQR: 25.7–46.5), and 38.25 mg (IQR: 31.5–58.5) in groups G2–G4, respectively. During the follow-up S-ketamine remained nearly the same between G4 and G3 (Table 3)

**Table 3.** Cumulative S-ketamine consumption during the study after the beginning of PACU care

| Time after PACU start | G1<br>(n=25) | G2<br>(n=25) | G3<br>(n=25) | G4<br>(n=25) |
| --- | --- | --- | --- | --- |
| 24 h |  |  |  |  |
| Mean (SD) | 0.0 (0.0) | 18.5 (7.7) | 37.4 (15.9) | 45.9 (24.2) |
| Median [IRQ] | 0.0 [0.0, 0.0] | 17.8 [12.0, 23.4] | 35.5 [25.7, 46.5] | 38.3 [31.5, 58.5] |
| 48 h |  |  |  |  |
| Mean (SD) | 0.0 (0.0) | 28.9 (16.8) | 57.7 (33.6) | 66.4 (38.4) |
| Median [IRQ] | 0.0 [0.0, 0.0] | 23.4 [17.0, 31.8] | 53.8 [35.0, 64.5] | 55.1 [39.0, 89.8] |
| 72 h |  |  |  |  |
| Mean (SD) | 0.0 (0.0) | 37.3 (24.1) | 66.4 (38.4) | 81.6 (55.0) |
| Median [IRQ] | 0.0 [0.0, 0.0] | 29.8 [22.0, 46.0] | 63.7 [45.9, 85.3] | 66.8 [42.6, 111] |

PACU, postoperative care unit, G1, Patient controlled intravenous oxycodone 1 mg/ml (n=25); G2, Patient controlled intravenous oxycodone 1 mg/ml+ ketamine 0.25 mg/ml (n=25); G3, Patient controlled intravenous oxycodone 1 mg/ml+ ketamine 0.5 mg/ml (n=25); G4, Patient controlled intravenous oxycodone 1 mg/ml+ ketamine 0.75 mg/ml (n=25).

### Acute pain measurements during postoperative care

There was a small yet statistically significant mean change in postoperative NRS measured at rest over the first 24 h between groups G4 and G1 (standardized effect size: 0.17, 95% CI: 0.013–0.32, *P =* 0.033), but not between group G3 and G1 (standardized effect size: −0.097, 95% CI: −0.25–0.059, *P =* 0.223) or G2 and G1 (standardized effect size −0.052, 95% CI: −0.21–0.10, *P =* 0.51) (Fig 3A). Age, sex, weight, chronic pain, and prior use of weak opioids or gabapentinoids had no effect on the results.

**Figure 3.**
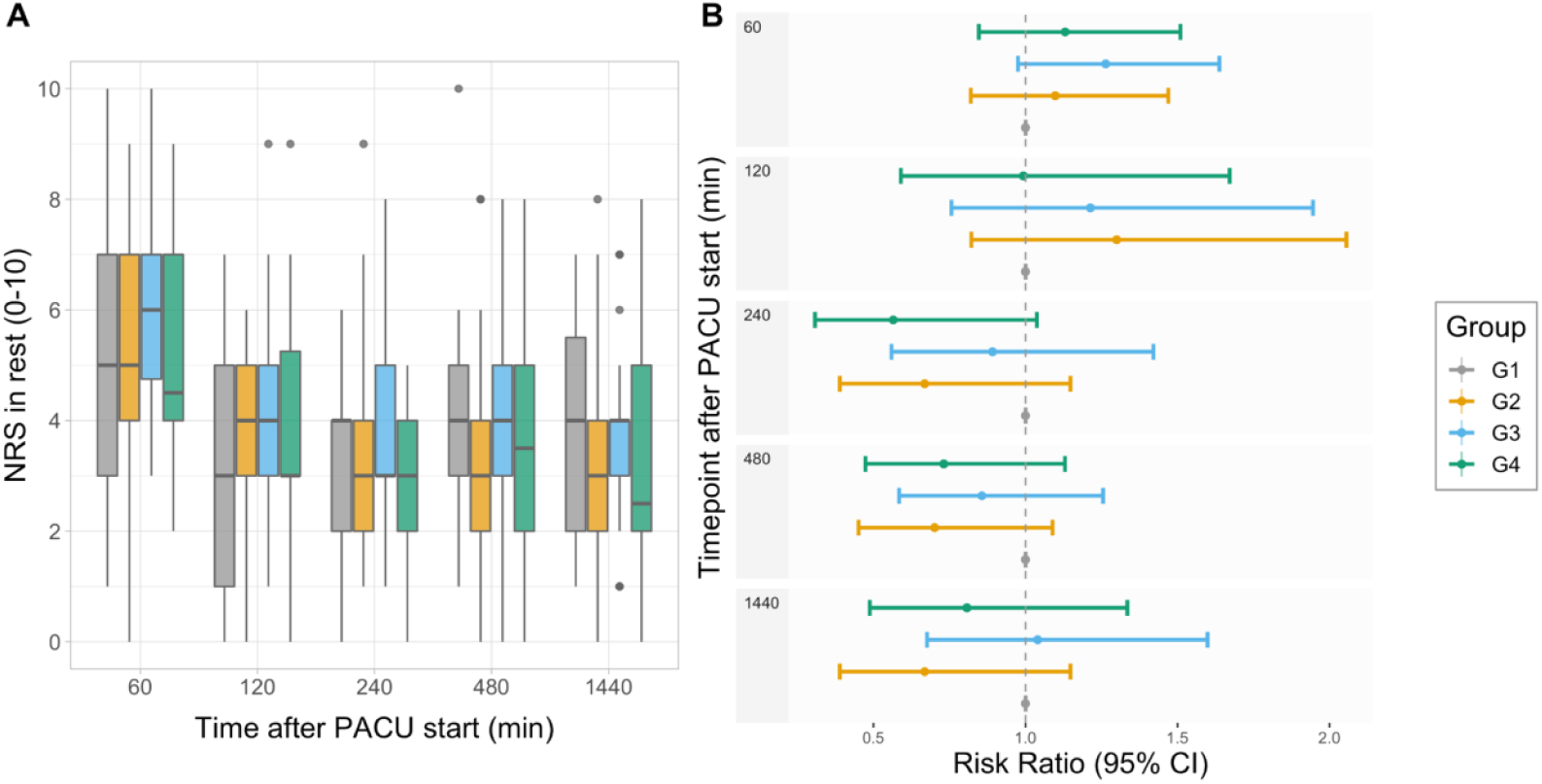
A) Patient-reported numerical rating scale (NRS) values at rest during the first 24 h in four patient-controlled analgesia (PCA) treatment groups G1-G4. Box plots show the median and 25–75th percentiles, and the whiskers indicate the minimum and maximum. (B) Risk ratios (95% CI) for reporting NRS value 4 or higher. PACU, postoperative care unit; CI, confidence interval.

To evaluate the effect of pain ratings as a surrogate for patient satisfaction, we analyzed the risk of having NRS >3 at rest at different timepoints during PACU treatment. For this purpose, NRS values were dichotomized depending on the pain level that patients experienced (NRS >3 or NRS ≤3). Although patient satisfaction, measured by patient-reported NRS was smaller in groups G2–G4 than group G1 at the end of PACU treatment, the finding was not statistically significant (Fig 3B).

### Postoperative sedation

At arrival to PACU, median (IQR) RASS was −1 (−2–0), and all patients were cooperative. RASS increased to 0 (−1 to 0) within the following 30 minutes. After 60 minutes, all patients were fully awake (median [IQR] RASS: 0 [0, 0]), and they remained fully awake until the end of the study. No significant differences in RASS scores were present between the groups during the 72-hour follow-up (Table 4).

**Table 4.** Opioid related adverse events during the study.

| Adverse effect |  | All | G1 | G2 | G3 | G4 |
| --- | --- | --- | --- | --- | --- | --- |
| Sedation (RASS-scale) |  |  |  |  |  |  |
| 0 min |  | -1 [-2, 0]<sup>1</sup> | -1 [-2, 0] | -1 [-3, 0] | -1 [-1, 0] | -1 [-2, 0] |
| 5 min |  | -1 [-2, 0] | -1 [-2, 0] | 0 [-1, 0] | 0 [0, -1] | -1 [-1, 0] |
| 30 min |  | 0 [0, 1] | 0 [0, -1] | 0 [-1, 0] | 0 [-1, 1] | 0 [-1, 0] |
| 60 min |  | 0 [0, 0] | 0 [0, -1] | 0 [0, 0] | 0 [0, 0] | 0 [0, 0] |
| 2 hr |  | 0 [0, 0] | 0 [0, 0] | 0 [0, 0] | 0 [0, 0] | 0 [0, 0] |
| 4 hr |  | 0 [0, 0] | 0 [0, 0] | 0 [0, 0] | 0 [0, 0] | 0 [0, 0] |
| 8 hr |  | 0 [0, 0] | 0 [0, 0] | 0 [0, 0] | 0 [0, 0] | 0 [0, 0] |
| 24 hr |  | 0 [0, 0] | 0 [0, 0] | 0 [0, 0] | 0 [0, 0] | 0 [0, 0] |
| PONV |  | n=100 | n=23 | n=25 | n=25 | n=25 |
| 24 hr | Yes (%) | 16 (16) | 2 (8) | 3 (12) | 6 (24) | 5 (22) |
| 48 hr | Yes (%) | 11 (11) | 2 (8) | 1 (4) | 5 (20) | 3 (13) |
| 72 hr | Yes (%) | 11 (11) | 2 (8) | 1 (4) | 5 (20) | 3 (13) |
| Pruritus |  | n=100 | n=25 | n=25 | n=25 | n=25 |
| 24 hr | Yes (%) | 16 (16) | 4 (16) | 7 (28) | 4 (16) | 1 (4) |
| 48 hr | Yes (%) | 10 (10) | 4 (16) | 4 (16) | 1 (4) | 1 (4) |
| 72 hr | Yes (%) | 7 (7) | 2 (8) | 1 (4) | 2 (8) | 2 (8) |
| Unpleasant dreams |  | n=100 | n=25 | n=25 | n=25 | n=25 |
| 24 hr | Yes (%) | 9 (9) | 2 (8) | 2 (8) | 2 (8) | 3 (12) |
| 48 hr | Yes (%) | 10 (10) | 4 (16) | 4 (16) | 1 (4) | 1 (4) |
| 72hr | Yes (%) | 7 (7) | 2 (8) | 1 (4) | 2 (8) | 2 (8) |

<sup>1</sup> RASS-scale given as median [interquartile range]

RASS, Richmond Agitation-Sedation Scale; PONV, postoperative nausea and vomiting; G1, Patient controlled intravenous oxycodone 1 mg/ml (n=25); G2, Patient controlled intravenous oxycodone 1 mg/ml+ ketamine 0.25 mg/ml (n=25); G3, Patient controlled intravenous oxycodone 1 mg/ml+ ketamine 0.5 mg/ml (n=25); G4, Patient controlled intravenous oxycodone 1 mg/ml+ ketamine 0.75 mg/ml (n=25).

### Opioid-related adverse effects

Opioid-related adverse events during the study are summarized in Table 4. Nausea and vomiting were the most frequent adverse events. At 24 h after PCA start, 16% of patients (n = 16) reported nausea. Increasing ketamine dose seemed to increase the incidence of PONV, but the changes were not statistically significant. Similarly, no differences were seen at the end of 48- and 72-h follow-up. We analyzed these adverse effects further with logistic regression. Our results indicate that preoperative weak opioid use increased the incidence of PONV significantly (odds ratio: 9.23, 95% CI: 1.4–75). Adjuvant ketamine in PCA had no effect (odds ratio: 3.9, 95% CI: 0.68–4.7) on PONV, and after changing to normal PCA with only oxycodone at 24 h, the incidence of PONV did not change (odds ratio: 0.53, 95% CI: 0.62–4.7).

At 24 h after PCA start, 16% of patients (n = 16) reported pruritus (Table 4). The PCA treatment groups did not show statistically significant differences at any of the three time points (24, 48, or 72 h after the start of PCA). Similarly, the incidence of pruritus did not change after changing to normal PCA with oxycodone only.

Nine patients (9%) reported nightmares or unpleasant dreams (Table 4). The groups showed no difference in this regard, and the incidence did not change after changing to normal PCA with oxycodone only. Age, sex, weight, chronic pain, and prior use of gabapentinoids had no effect on the results.

### Other adverse effects

There were no severe adverse effects. None of the patients developed respiratory insufficiency requiring invasive or non-invasive ventilation during the 72-h study period. No rescue medications were required during the study.

## Discussion

This randomized, double-blind, placebo-controlled study evaluated the effect of adding incremental doses of S-ketamine to oxycodone PCA in patients who underwent major lumbar spinal fusion surgery. Our results demonstrate that patients who received oxycodone:S-ketamine ratio 1:0.75 (a bolus containing oxycodone 1 mg + S-ketamine 0.75mg per ml) for postoperative analgesia consumed significantly less oxycodone postoperatively compared with participants who received lower S-ketamine doses or oxycodone alone. This oxycodone sparing effect was achieved with a mean 45.9 mg of S-ketamine during the first 24 h after lumbar spinal fusion surgery.

Additionally, there was a significant opioid sparing effect long after the S-ketamine was removed from PCA at 24 h postoperatively among patients who received oxycodone:S-ketamine ratio 1:0.75 compared with participants in other groups. This could be associated with the long-term antagonism of the NMDA-receptor or the recently proposed analgesic action of active metabolites [24].

A statistically significant effect in mean change in pain intensity at rest was seen in the group receiving 0.75 mg ml^−1^ S-ketamine in oxycodone PCA compared with patients receiving lower doses of ketamine or oxycodone alone. Cumulative oxycodone dose or adjunct S-ketamine did not significantly influence postoperative nausea and vomiting, which could be explained by the low cumulative consumption of oxycodone and ketamine in this group compared to others. Occurrence of pruritus, nightmares, or unpleasant dreams did not differ significantly among study groups.

Major spinal surgery is increasing in frequency and complexity. It is associated with severe postoperative pain [1], which is not easily amenable to regional anesthesia and requires a multimodal approach. This is a crowded field of clinical research, with many previous studies evaluating various single doses of ketamine with different opioids in many types of surgery. Yet, there is little previous data that assesses the effect of adjunct ketamine to an IV opioid-PCA exclusively after lumbar spinal fusion surgery in an opioid-naïve patient population. A recent review and meta-analysis evaluated the effect of perioperative ketamine for analgesia in spine surgery [25]. However, in most of the included trials, ketamine was administered intraoperatively followed by postoperative IV-PCA or ketamine was given both as a pre-incisional bolus and as a postoperative IV-PCA with a background infusion and a bolus on-demand [26,27]. This may confuse evaluation of the analgesic effect of IV-PCA administration of ketamine with an opioid. Additionally, the meta-analysis included patients undergoing lumbar microdiscectomy with postoperative opioid IV-PCA [28]. It could be anticipated that pain after lumbar microdiscectomy is less severe than after major lumbar spinal fusion surgery, and thus postoperative conditions are not comparable. Furthermore, one previous study was an open-label trial [29] and another had only female patients [30], making it difficult to generalize results.

Previous studies have shown an opioid-sparing effect of intravenous low-dose ketamine after spine surgery in opioid-dependent patients [31,32]. Additionally, Loftus (2010) and Nielsen (2017) showed that the benefit of ketamine increased with the amount of preoperatively administered opioids [31,32]. Recently Nielsen et al. (2019) showed that perioperative intravenous S-ketamine reduced analgesic use and pain and improved labor market attachment of opioid-dependent patients one year after spine surgery [33]. Another recent study further confirmed that postoperative low-dose ketamine infusion reduced hydromorphone requirements for the first 24 h after spinal fusion surgery in opioid-tolerant patients, but not in opioid-naïve patients [34]. However, it is recognized that previous opioid use alters pain processing, and these patients typically require higher opioid doses postoperatively. Thus, results of these studies cannot be directly applied to all patients.

Although the effect of adjunct ketamine in the opioid-tolerant patient population after lumbar fusion surgery is quite well established, the effect in the opioid-naïve population is less clear. Additionally, the optimal dose of adjunct ketamine in an opioid IV-PCA is unknown. Data indicating low incidence of ketamine-related adverse events (PONV and CNS adverse events) has been consistent in previous studies [11,12,35]. Likewise, our study showed that adjunct S-ketamine in oxycodone IV-PCA was well tolerated. There are several reports of ketamine’s beneficial effect on analgesia, opioid-sparing, and PONV in the postoperative period, without increasing the risk for hallucinations. However, the opioid:ketamine ratio in IV-PCA in previous studies was heterogeneous, ranging from 1:0.5 to 1:2.5, and these studies were also heterogeneous in regard to the opioid used, anesthesia methodology, type of surgery, patient population, and use of racemic- or S-ketamine [35,36].

Our study has limitations. Our study was designed to detect a difference in 24 h cumulative oxycodone consumption, which was the primary outcome. The beneficial effect of adjunct S-ketamine in an oxycodone IV-PCA in reducing the 24 h oxycodone consumption did not correlate with a reduction in opioid-related side effects, but this could be secondary to a lack of power. On the other hand, the strengths of our study include that the patient population was homogenous across study groups, consisting of adult men and women who underwent posterolateral lumbar spine fusion with bilateral transpedicular screw instrumentation.

Postoperative analgesic consumption as a surrogate measure for pain has been criticized on the basis of reports that it is skewed, where a minority of patients consume more than half of the postoperative analgesic. A recent study encouraged reporting categorized parameters as more clinically intuitive [37]. As discussed before, dichotomizing continuous measures leads to several problems [38]. Previously, analgesic consumption has been analyzed with point estimates or determining area under the curve, but both of these methods introduce considerable bias. Statisticians recommend using a model-based approaches, which has been used here. Therefore, both mean (SD) and median (IQR) have been reported. From a clinical perspective, it may be unrealistic to achieve a totally pain-free state after major surgical trauma following surgery such as instrumented lumbar spinal fusion. We think that IV-PCA enables the patient to titrate the opioid to reach a certain individual, tolerable pain intensity level. Therefore, we consider that changes in cumulative opioid consumption could serve as an acceptable surrogate for changes in postoperative pain. However, we also evaluated the outcome by using risk ratios for pain intensity NRS >3 as a surrogate for perioperative pain experience, and adjunct S-ketamine seemed to decrease the likelihood of pain exceeding NRS >3.

Our study adds new data considering the optimal dose of adjunct S-ketamine to an oxycodone IV-PCA after lumbar spinal fusion surgery. Because the oxycodone:S-ketamine ratio of 1:0.75 was not associated with increased adverse events, we suggest that future studies aimed at solving the optimal opioid:S-ketamine ratio evaluate that dose and higher, possibly with pharmacokinetic testing to characterize the dose-concentration-effect relationship.

In conclusion, IV-PCA containing adjunct S-ketamine with oxycodone at a ratio of 1:0.75 after major lumbar spinal fusion surgery is effective in decreasing the total oxycodone consumption at 24 h after surgery. This oxycodone:S-ketamine ratio is also well tolerated. As previous studies have mainly focused on intraoperative administration, even showing no analgesic effect of intraoperative S-ketamine in opioid-naïve patients [39], this finding adds new data to the feasibility of adjunct S-ketamine with oxycodone for postoperative pain management after major lumbar spinal fusion surgery.

## Data Availability

According to Finnish Medical Research Act (488/1999) data cannot be made available. The corresponding author gives further information on this matter.

## Acknowledgments

Eliisa Löyttyniemi, MSc, Institute of Biomedicine, University of Turku gave invaluable statistical help. The nursing staff of K- and T-operating units, TYKS ORTO, and neurosurgical wards at Turku University Hospital (Turku, Finland) and the staff of surgical ward 2 and the anesthesia nurses of the Department of Orthopaedics and Trauma Surgery at Töölö Hospital (Helsinki, Finland) are acknowledged for their support conducting the study. Sini Alanko, RN; Toni Broman, RN; Virva Kuusisto, RN; and Jaana Virtanen, RN are thanked for their help at the preoperative clinic. Tuula Eklund, RN; Ulla Simola, RN; and Saku Koskinen, RN are acknowledged for their generous help with the patient data system. Local hospital pharmacies in Turku and Helsinki University Hospitals are acknowledged, and ward pharmacists Anne Honkanen, Kira Honkanen, Elina Luoto, Leea Rissanen, Mari Saalasti and Reija Silvennoiovnen are thanked for fruitful collaboration.

## Notes

### Competing Interest Statement

The authors have declared no competing interest.

### Clinical Trial

NCT02994173

### Funding Statement

This work was supported by a governmental research grant from the Hospital District of South-West Finland, Finland (#13821)(T.I.S.).

### Author Declarations

Institutional review board of the Hospital District of Southwest Finland (number: 103/1800/2016) and the Finnish Medicines Agency (FIMEA, KL 135/2016).

### Summary of Updates

BACKGROUND: Spinal fusion surgery causes severe pain. Strong opioids, commonly used as postoperative analgesics, may have unwanted side effects. S-ketamine may be an effective analgesic adjuvant in opioid patient-controlled analgesia (PCA). However, the optimal adjunct S-ketamine dose to reduce postoperative opioid consumption is still unknown. METHODS: We randomized 107 patients at two tertiary hospitals in a double-blinded, placebo-controlled clinical trial of adults undergoing major lumbar spinal fusion surgery. Patients were randomly allocated to four groups in order to compare the effects of three different doses of adjunct S-ketamine (0.25, 0.5, and 0.75 mg ml^−1^) or placebo on postoperative analgesia in oxycodone PCA. Study drugs were administered for 24 hours postoperative after which oxycodone-PCA was continued for further 48 hours. Our primary outcome was cumulative oxycodone consumption at 24 hours after surgery. RESULTS: Of the 100 patients analyzed, patients receiving 0.75 mg ml^−1^ S-ketamine in oxycodone PCA needed 25% less oxycodone at 24 h postoperatively (61.2 mg) compared with patients receiving 0.5 mg ml^−1^ (74.7 mg) or 0.25 mg ml^−1^ (74.1 mg) S-ketamine in oxycodone or oxycodone alone (81.9 mg) (mean difference: −20.6 mg; 95% confidence interval [CI]: −41 to −0.20; P = 0.048). A beneficial effect in mean change of pain intensity at rest was seen in the group receiving 0.75 mg ml^−1^ S-ketamine in oxycodone PCA compared with patients receiving lower ketamine doses or oxycodone alone (standardized effect size: 0.17, 95% CI: 0.013-0.32, P = 0.033). The occurrence of adverse events was similar among the groups. CONCLUSIONS: Oxycodone PCA containing S-ketamine as an adjunct at a ratio of 1: 0.75 decreased cumulative oxycodone consumption at 24 h after major lumbar spinal fusion surgery without additional adverse effects.

